# Factors determining hemoglobin levels in vaginally delivered term newborns at public hospitals in Lusaka, Zambia

**DOI:** 10.1101/2024.08.11.24311826

**Authors:** Adenike Oluwakemi Ogah, Chrispin Mwando, Kenneth Chanda, Selia Nganjo

**Author notes:** **Corresponding Author:**. Phone contact.

## Abstract

**Background:** Limited data is available regarding the prevalence of neonatal anemia and its associated risk factors in areas with constrained resources.

**Subject and methods:** In a cross-sectional study, data pertaining to socio-demographic and clinical characteristics of 489 mother-singleton, term newborn pairs were consecutively collected from the admission wards of six public hospitals in Lusaka. The information was then analyzed to determine the prevalence of newborn anemia and its associated risk factors. Newborn and maternal anemia were defined as hemoglobin levels below 15g/dl and 11g/dl, respectively. The relationship between the variables was explored using Chi-square tests and a binary logistic regression model. The findings were reported in terms of p-values, odds ratios, and 95% confidence intervals.

**Results:** The prevalence of anemia and severe anemia in newborns was 72.4% and 2.5% respectively, while in mothers it was 30.5% and 14.7% respectively. Delayed cord clamping was performed in 71.4% of the deliveries, and 86.5% of newborns had their hemoglobin levels estimated between 4-6 hours after birth. Maternal pre-delivery hemoglobin data were obtained from the hospital records of 246 (49.7%) out of the 489 mothers in the study. The majority (63.4%) of maternal hemoglobin levels were determined more than 4 weeks before delivery, and this infrequent hemoglobin assessment was significantly associated with newborn anemia (p<0.001; OR 3.60; 95% CI 1.81, 7.14). Additionally, 11% of the 489 mothers had underlying medical conditions, which were also significantly associated with newborn anemia (p=0.019; OR=2.96; [95%CI 1.20, 7.32]). The top three maternal medical conditions were HIV (35.2%), hypertension (25.9%), and Hepatitis B virus infection (13%). Maternal age was significantly associated with newborn anemia, with teenage pregnancy posing the highest risk (93.8%; p<0.001; OR 5.68; [95%CI 1.94, 16.60]). Furthermore, primiparous (p<0.001; OR 5.46 [95% CI 2.02, 14.93]), para 2 (p=0.014; OR=2.11 [95% CI 1.16, 3.83]), and multiparous (p=0.009; OR=4.77; [95% CI 1.48, 15.35]) mothers were more likely to produce anemic newborns compared to other parity. Newborns born before 40 weeks gestation were 3.11 times (95% CI 1.75, 5.52) more likely to have anemia, p<0.001, compared to full-term babies. Normal birthweight babies were less likely to become anemic compared to low birthweight babies (p=0.003; OR=0.31 [95% CI 0.14, 0.68]).

**Conclusion:** Enhanced antenatal care for pregnant mothers in resource-limited settings is essential, with particular focus on maternal hemoglobin, nutrition, and medical conditions. Attention should also be given to teenage pregnancy, primiparous and multiparous mothers, as well as preterm and low birthweight babies, to prevent newborn anemia and consequently reduce infant morbidity and mortality.

## Background

Anemia is defined as a hematocrit (HCT) level of less than two standard deviations measured for a child’s age and gender.^1,2^ Anemia is defined in the newborn period using both gestational and postnatal age.^3^ The leading causes of anemia in newborns include blood loss, decreased red blood cell synthesis, accelerated red blood cell breakdown, low birth weight, and prematurity.^4,5,6^

Anemia’s clinical signs include poor feeding, rapid breathing, paleness, and a change in mental status due to poor systemic tissue perfusion.^7,8^ Tissue hypoxia, delayed brain development, stunted growth, chronic heart failure, infectious diseases such as HIV and hepatitis as a result of repeated blood product infusions, and eventually multiple organ failure are all complications of unrecognized and untreated anemia in infants.^9,10,11,12,13^

Globally, the prevalence of anemia during pregnancy is 42%, while newborn anemia in Sub-Saharan Africa is between 25 and 30%. In a prospective cohort study of 352 pregnant mothers in South western Uganda, Ngozi et al. found a 17% prevalence of newborn anemia based on umbilical cord blood and hemoglobin levels <13g/dl.^14^ The Uganda study identified maternal anemia, cesarean delivery, high maternal parity, and young maternal age as risk factors for neonatal anemia. In Ethiopia, 22% of pregnant women are anemic.^15,16^

Infants under six months of age are at significant risk of anemia due to their rapid growth and insufficient iron intake, as breast milk is poor in iron.^17^ As a result, they rely mostly on iron from intrauterine life.^18^ It is much more damaging when the pregnant mother, who is the primary source of fetal iron, is anemic.

Although newborn anemia can have serious consequences for neonates’ health and well-being throughout their lives, it receives little attention from healthcare practitioners and researchers in low-income nations. There have been insufficient studies undertaken on the prevalence of neonatal anemia in various parts of the world.^19^ There are no specific policies or guidelines for screening of newborn anemia in Africa. To the best of our knowledge, no other study has addressed this problem locally, and the 6 Public Hospitals involved in this study serve a large number of populations within and around the Central province of the country. Therefore, this study aimed to determine the prevalence of newborn anemia and associated factors in public Hospitals located in Lusaka disctrict of Zambia.

## Materials and methods

The methods employed in carrying out this study are discussed in this section.

### Study Design

This was a secondary analysis of a cross-sectional study.

### Study setting

The study was conducted at 6 public hospitals in Lusaka district: the Women and Newborn Hospital (the National Referral centre) and 5 first level hospitals.

Woman and Newborn Hospital is part of the University Teaching Hospitals. The University Teaching Hospital is the largest tertiary hospital in the country and is the highest national referral centre for nearly all patients in the country and those from all the 24 local clinics in the Lusaka district (including those from private health facilities). This hospital conducts at least 15,000 deliveries annually. It has a Neonatology Unit with a capacity to admit 60 newborns at once, but it often admits between 90 to 100 newborns, both inborn and out born. Additionally, the Kangaroo Mother Care unit can accommodate 26 neonates. Women and Newborn Hospital offers a range of obstetric and neonatal care, including labour and delivery services, family planning, postnatal care, neonatal intensive care, and support for premature or ill newborns.

The First Level Hospitals included Chawama, Chilenje, Chipata, Kanyama, and Matero hospitals. In the Zambian context, First Level Hospitals are akin to district hospitals and serve as referral centres within their respective constituencies. They are strategically located in the peri-urban areas of Lusaka, catering to the healthcare needs of the local population within their localities. In addition to the general medical and surgical services, the hospitals provide essential maternal and neonatal services, including maternity care and deliveries. Expectant mothers can access skilled birth attendants and receive necessary care during childbirth.

Matero General level hospital is the largest General hospital in Lusaka and is situated in the north-eastern part of Lusaka. It covers a high-density area. This facility had 3 qualified obstetricians and 103 supportive staff including qualified nurses and medical officers. The hospital had approximately 135 bed capacity and through the referral system, covered the majority of clinics in the north eastern region of Lusaka.

Chawama General level hospital is located in Chawama compound which is a high density area located in the south-western part of Lusaka. This hospital has a total for 2 qualified obstetricians that head the obstetrics and gynaecological department with approximately 23 medical staff.

Chilenje General Hospital is also located in a high density area catering to its neighbouring clinics. This facility has a total of 3 obstetricians, 2 senior house officers, 1 medical licentiate and 32 nurses.

Kanyama Level One Hospital has a high patient volume with an average monthly attendance of 23,400 and performs 35 – 50 deliveries daily. It serves a catchment population of 262,715 and is considered a high-volume site with low socioeconomic status

### Sampling Method

Six Research Assistants were recruited in each of the Public Hospitals. Sampling technique was consecutive, on first-come-first-serve basis until the desired sample size of 572 was obtained. Recruitment of participants in all the 6 Public Hospitals took place concurrently. The mother-newborn pair were recruited from the admission wards of the 6 public hospitals located in Lusaka District. The study was conducted over a 5-month period between 15th January to 3rd June 2024 at the 6 public hospitals as outlined in Table 1.

**Table 1.** Sample Distribution.

| Setting | Percentage | Sample size |
| --- | --- | --- |
| Women and Newborn Hospital | 40% | 229 |
| First level hospitals | 60% | 343 |
| --Chawama | 12.1% | 69 |
| --Chilenje | 12% | 68 |
| --Chipata | 12.1% | 69 |
| --Kanyama | 12.1% | 69 |
| --Matero | 12% | 68 |
| Total | 100% | 572 |

83 (14.5%) out of the 572 participants recruited, had incomplete hospital records, hence only data from 489 mother-newborn pairs were analysed.

### Data source and sample

The mother-newborn’s socio-demographic and clinical data were collected using structured questionnaire based-interviews that were created in English and translated to the local languages. Maternal body mass index (BMI) was calculated from measured weight (kg) and height (cm) using the standard method during the interview. Latest maternal Hb and date were obtained from the hospital file. The weight of the newborn baby was determined by midwives at birth. Timing of umbilical cord clamping after birth was recorded.^20^ Newborn Hemoglobin was determined with a portable hemoglobinometer, the HemoCue Hgb analyzer (HemoCueHb 201+, Sweden) according to standard guidelines within the first 24 hrs after birth. In this study, newborn anemia was defined as Hgb <15g/dL and maternal anemia as Hgb <11g/dL.^21,22^

### Statistical analysis

The data was manually cleaned, processed, checked for completeness and entered into Microsoft Excel. It was then exported into SPSS version 26 for analysis. After categorizing and defining the variables, a descriptive analysis was carried out for each of the independent variables and presented with numbers, frequencies and percentages. Chi test and Binary logistic regression analysis were used to assess the relationship between the newborn anemia and each independent variable. Multicollinearity and fitness of the model were checked. Factors with p-values <0.1 were included in the regression model. Odds ratio (OR), with a 95% confidence interval (CI) were computed. For all, statistical significance was declared at p-value <0.05. The reporting in this study were guided by the STROBE guidelines for observational studies.^23^

### Ethics

The Institutional Review Board of the University of Zambia (UNZABREC) approved original study titled ‘ASSESSING THE IMPACT OF DELAYED CORD CLAMPING ON NEONATAL HAEMOGLOBIN LEVELS IN VAGINAL DELIVERIES AT THE WOMEN AND NEWBORN HOSPITAL AND PUBLIC FIRST LEVEL HOSPITALS IN LUSAKA DISTRICT, ZAMBIA’ with reference number 4998-2024 was the source of the data for this article. Additional approvals from NHRA, WNH, and Lusaka Provincial Health Office were obtained before data could be collected. Further permission was obtained from Senior Medical Superintendent at each of the first level hospitals. The mother’s written informed consent and assent were obtained. Participants were assured of confidentiality as well as anonymity. The participants were informed that they were free to withdraw from the study without any negative consequences. All study documents were secured under a locked cabinet. This study posed no harm to the participants as it was an observational study. For newborns diagnosed with anemia, communication was established with the attending Doctor for further assessment and treatment. In order to protect the privacy and confidentiality of the participants, no personal identification such as name was collected. This study was carried out in accordance with the Helsinki Declaration. Results of the study will be made available to stakeholders to obtain information, which could be used for strategies to improve perinatal care.

## Results

The following are the results obtained from the study.

### Participants

The study targeted pregnant women giving birth at WNH-UTH and all 5 first level Hospitals in Lusaka district.

### Clinical characteristics of the newborns in the study

Of note, only 135 (27.6%) newborns out of the 489 recruited had normal hemoglobin levels of 15g/dl and above and 12 (2.5%) were severely anaemic with Hb <10g/dl, Table 2.

**Table 2:**
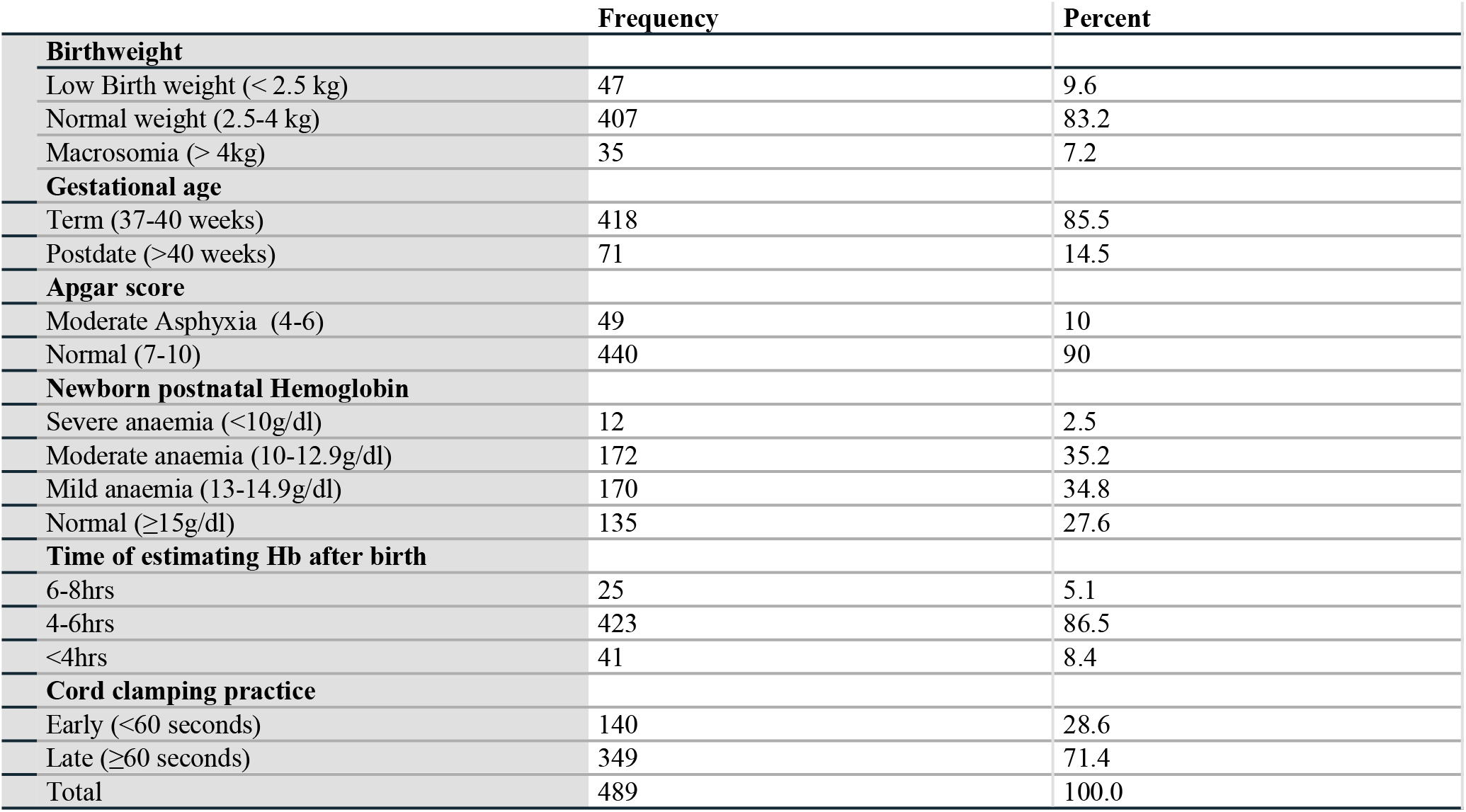
Clinical characteristics of newborns in the study, n=489.

A higher percentage of the mothers were young (>25-35years of age, 43.1%), overweight (47.9%), of secondary school education (50.3%), primiparous (40.7%) with no underlying medical condition (89.0%) and have never had a miscarraige (81.0%). Of note, the teenagers had a higher percentage (9.4%) of those with low BMI, compared to other age groups, p<0.001 (OR 3.84; 95%CI 1.03,14.36), Table 3.

**Table 3:** Maternal characteristics in the study, n=489.

| Maternal Characteristics | Frequency | Percent |
| --- | --- | --- |
| **Age** |  |  |
| Teenage (< 18 years) | 32 | 6.5 |
| Very young (18-25 years) | 209 | 42.7 |
| Young (>25-35 years) | 211 | 43.1 |
| Elderly (>35 years) | 37 | 7.6 |
| **BMI (kg/m<sup>2</sup>)** |  |  |
| Wasted (<18.5) | 15 | 3.1 |
| Normal (18.5-24.9) | 174 | 35.6 |
| Overweight (25-29.9) | 234 | 47.9 |
| Obese ($>30$) | 66 | 13.5 |
| **Education** |  |  |
| None | 31 | 6.3 |
| Primary | 144 | 29.4 |
| Secondary | 246 | 50.3 |
| Tertiary | 68 | 13.9 |
| **Maternal Medical (present or absent)** |  |  |
| Presence of underlying medical condition | 54 | 11.0 |
| Absence of underlying medical condition | 435 | 89.0 |
| **Parity (delivery)** |  |  |
| 1 | 199 | 40.7 |
| 2 | 108 | 22.1 |
| 3 | 100 | 20.4 |
| 4 | 64 | 13.1 |
| >4 | 18 | 3.7 |
| **Miscarraige (Yes/No)** |  |  |
| Yes | 93 | 19.0 |
| No | 396 | 81.0 |
| Total | 489 | 100.0 |

Proportion of women with HIV infection was the highest amongst the maternal medical conditions in the study (19/54=35.2%), followed by hypertension (14/54=25.9%) and Hepatitis B virus infection (13.0%), Table 4.

**Table 4:** Maternal medical diagnosis in the study, n=54.

| Maternal Medical Diagnosis | Frequency | Percent |
| --- | --- | --- |
| Diabetes Mellitus | 3 | 5.6 |
| Hepatitis B Virus infection | 7 | 13 |
| Heart Disease | 3 | 5.6 |
| HIV infection | 19 | 35.2 |
| Hypertension | 14 | 25.9 |
| Pregnancy Induced Hypertension | 4 | 7.4 |
| Sickle Cell Disease | 4 | 7.4 |
| Total | 54 | 100.0 |

Only 246 out of the 489 mothers in the study had a record of hemoglobin in their hospital files. 75 (30.5%) of these 246 mothers were anemic. 11 (14.7%) of the 75 anemic mothers had severe anemia. Majority, (156, 63.4%) of these 246 mothers had their last hemoglobin estimate performed more than 4 weeks before delivery. Of the 75 mothers, who were anemic, 59 (78.7%) of them had anemic newborns; compared to 125 (73.1%) of the 171 mothers that were not anemic, p=0.355; OR =1.36 (95% CI 0.71, 2.59). Majority of the 156 mothers, who had their pre-delivery hemoglobin performed more than 4 weeks prior (127, 81.4%) had anemic newborns, compared to only 28 (54.9%) out of the 51 mothers who had their pre-delivery hemoglobin performed <1 week before delivery (p<0.001; OR 3.60; 95%CI 1.81, 7.14), Table 5.

**Table 5:**
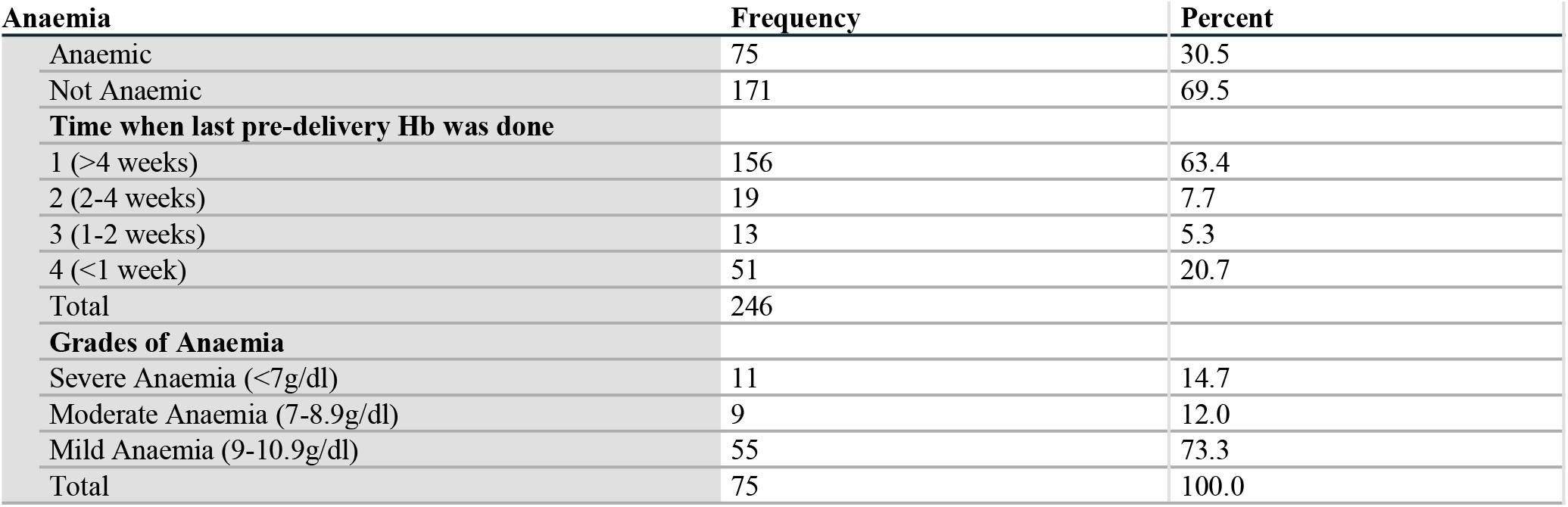
Maternal Hemoglobin levels in the study, n=246.

Univariate analysis was done to investigate the relationships between the independent variables and newborn anemia. Birthweight had no significant relationship with newborn hemoglobin. The early term newborns were 3.31 (95% CI:1.98, 5.56) times more likely to be anemic compared with the postdate newborns (p<0.001). Early cord clamping was more likely to result in newborn anemia, compared to delayed cord clamping (OR=1.34; 95% CI 0.85, 2.11), however, this was not significant, p=0.206. Maternal medical illness was signifcantly associated with neonatal anemia, p=0.011; OR=2.80 (95% CI 1.23, 6.36). Though, babies sampled more than 6hrs after birth, were more likely to be anemic (20 out of 25 [80%]) compared to those sampled within 4 hrs after birth (25 out of 41 [61%]); OR 2.56 (95% CI 0.80, 8.20), this relationship was not significant at p=0.175. Maternal age was significantly associated with newborn anemia (p<0.001). Teenage mothers (30 out of 32, 93.8%) were 5.68 times (95% CI 1.94, 16.60) more likely to have anemic babies, followed by the elderly mothers (33 out of 37, 89.2%). The young mothers were the least likely to have anemic newborns (125 out of 211, 59.2%). Parity was also significantly associated with newborn anemia, p<0.001. The Para 4 mothers (53 out of 64, 82.8%) were 6.02 times (95%CI 1.94, 18.87) more likely to have anemic newborns, followed by the primips (162 out of 199, 81.4%; OR 5.46 [95% CI 2.02, 14.93]), Table 6.

**Table 6:** Crosstabulation of factors contributing to neonatal postnatal hemoglobin in the study.

|  |  | Postnatal Neonatal Hb |  | Total |
| --- | --- | --- | --- | --- |
|  |  | Anaemic (%) | Not anaemic (%) |  |
| **Birthweight** | p-value=0.660 |  |  |  |
| Low Birth weight |  | 32 (68.1) | 15 (31.9) | 47 |
| Normal Birth weight |  | 298 (73.2) | 109 (26.8) | 407 |
| High Birth weight |  | 24 (68.6) | 11 (31.4) | 35 |
| **Gestational age** | p.value<0.001; OR=3.31 |  |  |  |
| Term |  | 319 (76.3) | 99 (23.7) | 418 |
| Postdate |  | 35 (49.3) | 36 (50.7%) | 71 |
| **Apgar score** | p.value=0.607; OR =1.20 |  |  |  |
| Moderate Asphyxia |  | 37 (75.5) | 12 (24.5) | 49 |
| Normal Apgar score |  | 317 (72.0) | 123 (28) | 440 |
| **Time of estimating newborn's Hb** | p.value= 0.175 |  |  |  |
| 6-8hrs |  | 20 (80.0) | 5 (20.0) | 25 |
| 4-6hrs |  | 309 (73.0) | 114 (27.0) | 423 |
| <4hrs |  | 25 (61.0) | 16 (39.0) | 41 |
| **Cord clamping** | p=0.206; OR=1.34 |  |  |  |
| Early |  | 107 (76.4) | 33 (23.6) | 140 |
| Late |  | 247 (70.8) | 102 (29.2) | 349 |
| **Maternal age (years)** | p<001 |  |  |  |
| Teenage (<18) |  | 30 (93.8) | 2 (6.3) | 32 |
| Very young (18-25) |  | 166 (79.4) | 43 (20.6) | 209 |
| Young (>25-35) |  | 125 (59.2) | 86 (40.8) | 211 |
| Elderly (>35) |  | 33 (89.2) | 4 (10.8) | 37 |
| **Maternal BMI** | p=0.452 |  |  |  |
| Wasted (<18.5) |  | 12 (80.0) | 3 (20.0) | 15 |
| Normal (18.5-24.9) |  | 120 (69.0) | 54 (31.0) | 174 |
| Overweight (25-29.9) |  | 176 (75.2) | 58 (24.8) | 234 |
| Obese (>30) |  | 46 (69.7) | 20 (30.3) | 66 |
| **Maternal Education** | p=0.072 |  |  |  |
| None |  | 28 (90.3) | 3 (9.7) | 31 |
| Primary |  | 108 (75.0) | 36 (25.0) | 144 |
| Secondary |  | 172 (69.9) | 74 (30.1) | 246 |
| Tertiary |  | 46 (67.6) | 22 (32.4) | 68 |
| **Maternal Medical illness** | p=0.011; OR= 2.80 |  |  |  |
| Yes |  | 47 (87.0) | 7 (13.0) | 54 |
| No |  | 307 (70.6) | 128 (29.4) | 435 |
| **Parity (# of deliveries)** | p<0.001 |  |  |  |
| 1 |  | 162 (81.4) | 37 (18.6) | 199 |
| 2 |  | 65 (60.2) | 43 (39.8) | 108 |
| 3 |  | 66 (66.0) | 34 (34.0) | 100 |
| 4 |  | 53 (82.8%) | 11 (17.2%) | 64 |
| >4 |  | 8 (44.4) | 10 (55.6) | 18 |
| **Miscarraige** | p=0.733; OR=0.92 |  |  |  |
| Yes |  | 66 (71.0) | 27 (29.0) | 93 |
| No |  | 288 (72.7) | 108 (27.3) | 396 |
| **Total** | **Count** | **354(72.4)** | **135(27.6)** | **489** |

**Table 7:** Binary Logistic regression analysis of variables determining newborn anemia, n=489.

| Step 1<sup>a</sup> |  | B | S.E. | Wald | df | p-value | OR | 95% C.I.for OR |  |
| --- | --- | --- | --- | --- | --- | --- | --- | --- | --- |
|  |  |  |  |  |  |  |  | Lower | Upper |
|  | Low birthweight |  |  | 8.80 | 2 | 0.012 |  |  |  |
|  | Normal birthweight | -1.180 | 0.40 | 8.55 | 1 | 0.003 | 0.307 | 0.14 | 0.68 |
|  | High birthweight | -.712 | 0.55 | 1.70 | 1 | 0.192 | 0.491 | 0.17 | 1.43 |
|  | Term gestation | 1.135 | 0.29 | 15.07 | 1 | <0.001 | 3.110 | 1.75 | 5.52 |
|  | Normal Apgar score | 0.543 | 0.41 | 1.74 | 1 | 0.187 | 1.720 | 0.77 | 3.85 |
|  | Delayed clamping | 0.357 | 0.26 | 1.95 | 1 | 0.162 | 1.429 | 0.87 | 2.36 |
|  | Teenager |  |  | 19.26 | 3 | <0.001 |  |  |  |
|  | Very young | 1.17 | 0.78 | 2.26 | 1 | 0.133 | 3.21 | 0.70 | 14.66 |
|  | Young | 2.07 | 0.79 | 6.97 | 1 | 0.008 | 7.94 | 1.71 | 36.96 |
|  | Elderly | 0.68 | 1.00 | 0.47 | 1 | 0.493 | 1.98 | 0.28 | 13.92 |
|  | Presence of Maternal medical condition | 1.09 | 0.46 | 5.55 | 1 | 0.019 | 2.96 | 1.20 | 7.32 |
|  | Parity 1 |  |  | 16.09 | 4 | 0.003 |  |  |  |
|  | Parity 2 | 0.75 | 0.31 | 5.98 | 1 | 0.014 | 2.11 | 1.16 | 3.83 |
|  | Parity 3 | 0.19 | 0.32 | 0.36 | 1 | 0.547 | 1.21 | 0.65 | 2.27 |
|  | Parity 4 | -0.39 | 0.43 | 0.81 | 1 | 0.367 | 0.68 | 0.29 | 1.58 |
|  | Parity>4 | 1.56 | 0.60 | 6.87 | 1 | 0.009 | 4.77 | 1.48 | 15.35 |
|  | Constant | -3.68 | 0.98 | 14.19 | 1 | 0.000 | 0.03 |  |  |

a. Variable(s) entered on step 1: Birthweight, Gest.2.age, Apgar2score, cclamp, Mat.age.cd, Med.Mat, Parity.

In the multivariate analysis using binary logistic regression model, the low birthweight, the term baby, teenage pregnancy, presence of maternal medical condition, Parity 1, 2 and the multiparous mothers remained significantly associated with newborn anemia.

## Discussion

The study examined the prevalence of anemia and its associated risk factors in 489 term, vaginally born newborns within the first 24 hours of life. It revealed a considerably higher rate of newborn anemia at 72.4% than the 29.1% prevalence reported in a facility-based study of 278 term babies in Ethiopia.^24^. The variation in the prevalence of anemia in newborns can be attributed to the utilization of different definitions of newborn anemia in the two studies. Specifically, the Ethiopian study employed a hemoglobin cut-off point of 13.5g/dl, whereas the Zambian study used a higher cut-off point of <15g/dl. Furthermore, the rate of low birth weight (LBW) in the Ethiopian study was notably lower at 2.2% compared to the 9.6% reported in the Zambian study, low birthweight being a known cause of newborn anemia. In the Ethiopian study, delayed cord clamping (1-3 minutes) was uniformly implemented for all deliveries, with birth umbilical cord blood being sampled for hemoglobin assessment. On the other hand, the Zambian study indicated that early cord clamping was executed in 28.6% of deliveries, and newborn hemoglobin levels were evaluated from foot heel capillary samples within 8 hours post-delivery. Furthermore, only 8.4% of blood sampling took place within 4 hours of life in the Zambian study. Notably, maternal anemia was less prevalent at 24.1% in the Ethiopian study compared to the 30.5% reported in the Zambian study. Additionally, the mothers’ mean age (26.21 years, sd=5.73) and nutritional status in the Zambian study were comparable to those in the Ethiopian study (mean 26 years, sd=4.4).

The prevalence of newborn anemia in the present study surpassed that of Rio de Janeiro, Brazil (32.6%), Lagos, Nigeria (35%), and Gondar, Ethiopia (25%). ^25,26,27^ However, the observed high rate of newborn anemia aligns with figures reported in Ghana (57.3%), Benin (61.1%), and southern Nigeria (65.6%).^28,29,30^

Although the prevalence of maternal anemia in the current Zambian study (30.5%) surpassed the 24.1% documented in the Ethiopian study, there was no significant association between maternal anemia and newborn anemia, in contrast to the findings in the Ethiopian and Ugandan studies. This disparity may be attributed to the fact that hemoglobin records were accessible for only 49.7% of the 489 sampled mothers in the Zambian study. However, the timing of pre-delivery maternal hemoglobin determination was significantly linked to newborn anemia in this Zambian study, with mothers whose hemoglobin levels were infrequently assessed during pregnancy showing an increased propensity to give birth to anemic newborns.

As the fetus receives nutrients, including iron and folate, from the mother, depleted maternal iron stores can prevent the fetus from accumulating sufficient iron, resulting in decreased fetal iron stores and reduced fetal hemoglobin levels.^31,32,33,34,35^ The physiological changes and metabolic demands of pregnancy result in an increased need for iron during pregnancy,^36^ predisposing women to maternal anemia. Maternal anemia induces adaptations in placental and fetal physiology, potentially leading to pregnancy and childbirth complications such as low birth weight, neurodevelopmental disorders, and premature delivery.^37, 38,39,40,41^

Maternal age was not found to be a significant determinant of newborn hemoglobin levels in either the Uganda or Ethiopia studies. However, in the Zambian study, teenage pregnancy showed a particular association with newborn anemia. Teenagers in the current study also displayed a higher likelihood of being malnourished compared to other age groups. While the Uganda study did not find a statistically significant association, it noted that younger mothers were more likely to have anemic newborns. Regarding parity, it was deemed significant in both the Zambian and Uganda studies. In the Zambian study, both primiparous and grandmultiparous mothers were significantly associated with newborn anemia. Likewise, in the Uganda study, higher maternal parity correlated with an increased likelihood of newborn anemia. Maternal medical conditions, particularly HIV, were identified as significant contributors to newborn anemia in this study. This study underscores the significance of quality antenatal care in ensuring the birth of full-term and normal birth weight babies, thus reducing the likelihood of postnatal anemia.

### Limitations

The available data lacked specific details concerning antenatal care and the sex of the newborn, factors that could potentially influence newborn hemoglobin levels. Additionally, the authors acknowledged the absence of a global consensus on standard cut-off points for newborn hemoglobin, as evidenced by the various definitions of newborn anemia used in different studies.

### Strength

This study, conducted across multiple centers, yielded valuable insights into antenatal care practices and newborn anemia in resource-limited settings, addressing a gap in the current literature. Additionally, while previous studies relied on umbilical cord-blood samples, the present study utilized peripheral blood collected from the heel site to estimate postnatal newborn hemoglobin levels, offering a more accurate depiction of the newborn hemoglobin profile.

## Conclusion ad Recommendations

Enhanced antenatal care for pregnant mothers in resource-limited settings should place greater emphasis on maternal hemoglobin levels, nutrition, and medical conditions. Additionally, specific attention should be directed towards teenage pregnancy, primiparous and multiparous mothers, as well as preterm and low birthweight babies, with the aim of preventing newborn anemia and subsequently reducing infant morbidity and mortality.

## Data Availability

All data produced in the present study are available upon reasonable request to the authors

## Author Contributions

The corresponding author (Dr Adenike Oluwakemi Ogah) co-supervised the study and the original dissertation write-up, conducted the secondary data analysis, interpreted the results and drafted this manuscript. Dr Chrispin Mwando, conceived the study title, collected the data and drafted the original dissertation. Dr Kenneth Chanda supervised the study and original dissertation write-up. Dr Selia Nganjo co-supervised the study and original dissertation write-up. All the authors contributed to the intellectual content of this manuscript and final editing of the article.

## Acknowledgements

The authors are extremely grateful to the participants involved in this study, to the staff of each of the Public Hospitals in Lusaka that contributed to this study and to the research team.

